# Risk, clinical course and outcome of ischemic stroke in patients hospitalized with COVID-19: a multicenter cohort study

**DOI:** 10.1101/2021.03.11.21253189

**Authors:** Wouter M. Sluis, Marijke Linschoten, Julie E. Buijs, J. Matthijs Biesbroek, Heleen M. den Hertog, Tessa Ribbers, Dennis Nieuwkamp, Reinier C. van Houwelingen, Andreas Dias, Ingeborg W.M. van Uden, Joost P. Kerklaan, H. Paul Bienfait, Sarah E. Vermeer, Sonja W. de Jong, Mariam Ali, Marieke J.H. Wermer, Marieke T. de Graaf, Paul J.A.M. Brouwers, Folkert W. Asselbergs, L. Jaap Kappelle, H. Bart van der Worp, Annemijn M. Algra, on behalf of the CAPACITY-COVID collaborative consortium

## Abstract

**Background and purpose:** The frequency of ischemic stroke in patients with COVID-19 varies in the current literature, and risk factors are unknown. We assessed the incidence, risk factors, and outcomes of acute ischemic stroke in hospitalized patients with COVID-19.

**Methods:** We included patients with a laboratory confirmed SARS-CoV-2 infection admitted in 16 hospitals participating in the international CAPACITY-COVID registry between March 1^st^ and August 1^st^, 2020. Patients were screened for the occurrence of acute ischemic stroke. We calculated the cumulative incidence of ischemic stroke and compared risk factors, cardiovascular complications, and in-hospital mortality in patients with and without ischemic stroke.

**Results:** We included 2147 patients with COVID-19, of whom 586 (27.3%) needed treatment at an intensive care unit (ICU). Thirty-eight patients (1.8%) had an ischemic stroke. Patients with stroke were older, but did not differ in sex or cardiovascular risk factors. Median time between onset of COVID-19 symptoms and diagnosis of stroke was two weeks. The incidence of ischemic stroke was higher among patients who were treated at an ICU (16/586; 2.7% versus 22/1561; 1.4%; p=0.039). Pulmonary embolism was more common in patients with (8/38; 21.1%) than in those without stroke (160/2109; 7.6%; adjusted RR: 2.08; 95%CI:1.52-2.84). Twenty-seven patients with ischemic stroke (71.1%) died during admission or were functional dependent at discharge and in-hospital mortality. Patients with ischemic stroke were at a higher risk of in-hospital mortality (adjusted RR 1.56; 95%CI:1.13-2.15) than patients without stroke.

**Conclusions:** In this multicenter cohort study, the cumulative incidence of acute ischemic stroke in hospitalized patients with COVID-19 was approximately 2%, with a higher risk in patients treated at an ICU. The majority of stroke patients had a poor outcome. The association between ischemic stroke and pulmonary embolism warrants further investigation.

## Introduction

Coronavirus disease 2019 (COVID-19) is a pandemic that has affected millions of people worldwide. Although respiratory symptoms are the predominant feature of the disease, the clinical course of COVID-19 may be complicated by venous and arterial thromboembolic events.^1, 2^ Pulmonary embolism accounts for the majority of these events, but other cardiovascular complications, including ischemic stroke, have also been reported. In contrast to some of the early reports suggesting an increased risk of acute ischemic stroke among patients hospitalized with COVID-19, results from later reports are less consistent. The occurrence of ischemic stroke varied greatly, ranging from 0.01% to 6.9%.^3-17^ This may be explained in part by differences in study design, size, case-findings methods, and settings. Studies that reported clinical details have suggested an increased severity of stroke symptoms, a higher incidence of cryptogenic strokes, and a worse outcome,^3, 4, 18^ including higher in-hospital mortality rates,^3-5^ in patients with COVID-19 than in those without. Nevertheless, large cohort studies reporting data on stroke severity, etiology, and outcome are still limited, probably due to the fact that ischemic stroke was often not reported as primary outcome and strokes were not assessed by neurologists. In addition, little data are available on the relationship between ischemic stroke and the occurrence of other cardiovascular complications in patients with COVID-19. To improve our understanding of the relation between COVID-19 and acute ischemic stroke, we assessed risk factors, time course, hospital setting, stroke severity, the relationship with other cardiovascular complications, and outcomes of ischemic stroke in patients hospitalized with COVID-19 during the first wave of the pandemic across 16 centers in the Netherlands.

## Methods

### Study design

This study was conducted within the CAPACITY-COVID international patient registry (www.capacity-covid-eu;NCT04325412). Within this registry, data generated during routine clinical care was collected in a standardized manner. Details regarding CAPACITY-COVID have been outlined in detail elsewhere.^12^ In short, CAPACITY-COVID extended the case report form (CRF) of the International Severe Acute Respiratory and emerging Infection Consortium (ISARIC) to collect in-depth information on cardiovascular history, the use of cardiovascular medication, and cardiac and thromboembolic events of patients hospitalized with COVID-19. STROCORONA was incorporated as a substudy within CAPACITY-COVID, to obtain additional detailed information on neurovascular history and the occurrence of acute ischemic stroke during hospitalisation, including data on vascular risk factors, etiology, severity, and outcome. Sixteen Dutch hospitals participated in STROCORONA. Ethical approval was obtained in all participating hospitals and the necessity of a consent procedure was determined conform local regulations. The majority of participating sites had an opt-out approach, were patients received written information about the study during hospitalisation.^19^ The data used for this study, including de-identified individual participant data and a data dictionary defining each field or variable within the dataset, can be made available upon reasonable request to the data access committee of CAPACITY-COVID. These data will be made available following publication of this work. A data sharing agreement must be signed before any data are shared.

### Study population and data collection

We included adult patients with a laboratory-confirmed SARS-CoV-2 infection (determined by a positive polymerase chain reaction (PCR) test result from a nasopharyngeal swab) who were admitted to a hospital during the first wave of the pandemic in the Netherlands (from March 1^st^ to August 1^st^, 2020). Patients who were suspected of COVID-19 but did not have a PCR-proven COVID-19 diagnosis were excluded. We retrieved data on demographics, comorbidities, pre-hospital medication, the need of mechanical ventilation, treatment at a high-dependency or intensive care unit (ICU) at any time during admission, in-hospital mortality, and the occurrence of other cardiac or thromboembolic complications: deep vein thrombosis (DVT), pulmonary embolism (PE), acute coronary syndrome (ACS), endocarditis, and new-onset atrial fibrillation (AF). Outcome definitions of in-hospital complications have been reported previously.^19^ For STROCORONA, patient files of all eligible cases were systematically screened and scored by neurologists or other physicians with experience in stroke research to identify acute ischemic stroke during hospitalisation. In addition, data on prior neurovascular diseases, including transient ischemic attack (TIA), ischemic stroke, intracerebral hemorrhage, subarachnoid hemorrhage, or vascular dementia, were collected. ‘Ischemic stroke’ was defined as a sudden onset of focal neurological signs originating from the brain or retina that persists for more than 24 hours or until death, confirmed with neuroimaging demonstrating either infarction in the corresponding vascular territory or absence of another apparent cause.^20^ We graded stroke severity at the time of diagnosis with the National Institutes of Health Stroke Scale (NIHSS; range 0 to 42, with higher scores indicating more severe neurological deficits) and collected data on acute stroke treatment (intravenous thrombolysis (IVT), endovascular treatment (EVT) and antithrombotic treatment), timing (median time between onset of COVID-19 symptoms and stroke diagnosis), and imaging findings (vascular territory, intracranial large vessel occlusion). We classified stroke etiology with the Trial of Org 10172 in Acute Stroke Treatment (TOAST) criteria and scored stroke outcome at discharge with the modified Rankin Scale (mRS).^21^

### Statistical analysis

Baseline characteristics are summarized with descriptive statistics as median (interquartile range; IQR), mean (standard deviation; SD), or frequencies (proportions) where appropriate. We did not impute missing data (details provided in Supplementary Material, please see https://www.ahajournals.org/journal/str). We calculated the cumulative incidence of ischemic stroke with corresponding 95% confidence intervals (95% CI) and stratified according to age and sex. Because three hospitals that participated in STROCORONA excluded patients that had no cardiovascular risk factors or were not consulted by a cardiologist during admission, we performed a sensitivity analysis excluding these three centers. We compared the occurrence of other cardiovascular complications and in-hospital mortality between patients with and without ischemic stroke with χ^2^ or Student’s t tests as appropriate and calculated risk ratios (RR) and 95% (CI) with Poisson regression.^22^ We adjusted RRs for age, sex, and treatment on an ICU. For stroke outcome, we calculated the proportion of ischemic stroke patients with an unfavorable outcome (defined as death or dependency (mRS of ≥3)) at discharge.

### Results

We included a total of 2147 patients in STROCORONA (Figure 1 of the Supplementary Material, please see https://www.ahajournals.org/journal/str). Table 1 shows the baseline characteristics. The median age was 70.0 years (IQR: 59.0-77.0), about one-third of the patients was female (769; 35.8%) and cardiovascular risk factors and comorbidities were common. Of all admitted patients, 586 (27.3%) received treatment at an ICU and 521 (24.3%) patients received mechanical ventilation. In general, patients treated at an ICU were younger and had fewer comorbidities (Table 2 of the Supplement). Acute ischemic stroke occurred in 38 of 2147 (1.8%; 95%CI: 1.3%-2.4%) patients (Table 2). These patients were older than patients without ischemic stroke and had a lower BMI, but did not differ in terms of sex, cardiovascular comorbidities, and pre-hospital medication (Table 1; please see also Table 3 in the Supplementary Material, https://www.ahajournals.org/journal/str). Also after stratification by age we did not find differences in cardiovascular risk factors between patients with and without ischemic stroke (Supplementary Table 4). In a sensitivity analysis excluding three hospitals that excluded patients without cardiovascular risk factors or cardiologist consultation during admission, baseline characteristics and cumulative stroke incidence were similar (Supplementary Table 5). The median time between onset of COVID-19 symptoms and stroke diagnosis was 14 days (IQR: 9-25 days) for all patients, 23 days (IQR: 13-29) for patients who received ICU treatment and 10 days (IQR: 3-18) for patients treated on a general ward only (p=0.031; Figure 1 and Table 3). Stroke symptoms occurred as presenting complaint in 4 (10.5%) patients. The cumulative incidence of acute ischemic stroke was 2.7% in patients who were treated at an ICU (16/586; 95% CI: 1.7%-4.4%) and 1.4% in patients who received treatment on a general ward alone (22/1561; 95% CI: 0.9%-2.1%; p=0.039). Age- and sex stratified cumulative incidence are given in Table 2 and details about stroke severity, subtype, imaging, acute treatment and outcome in Table 3. Stroke patients treated at an ICU were younger than those treated at a general ward only (ICU: 63.4 years (SD: 15.2); general ward: 79.2 (SD: 8.1); p<0.001), frequently had other thromboembolic events (ICU: 8/16 (50%); general ward: 2/22 (9.1%); p=0.020), and had more severe strokes (ICU: median NIHSS 22.0; IQR: 3.8-30.0; general ward: 5.0; IQR: 2.8-17.5; p=0.050; Table 3). Eighteen patients (47.4%) had a stroke of undetermined etiology, however in 6 (33.3%) the diagnostic workup was incomplete due to early withdrawal of care. Differences between patients with cryptogenic stroke and those with a stroke of known etiology are summarized in the Supplement (Table 6). The occurrence of other cardiovascular complications in patients with and without ischemic stroke is given in the Supplementary Material (Table 7, please see https://www.ahajournals.org/journal/str). Pulmonary embolism was more common in patients with (8/38 (21.1%)) than in patients without ischemic stroke (160/2109 (7.6%); p=0.002), also after adjustment for age, sex, and treatment on an ICU (aRR 2.08; 95% CI: 1.52-2.84). The median time between onset of COVID-19 symptoms and occurrence of PE was 18 days (IQR: 12-25 days) for all patients, 19 days (IQR: 12-26) for patients who received ICU treatment and 14 days (IQR: 8-21) for patients treated on a general ward only (p=0.04). In 5/8 (62.5%) patients with PE and acute ischemic stroke, PE was diagnosed before ischemic stroke. Three-quarters of the patients with ischemic stroke (27/38 (71.1%)) had an mRS of 3 or more (dead or dependent) at discharge (Figure 2). Patients with ischemic stroke were at a higher risk of in-hospital mortality (adjusted RR 1.56; 95% CI: 1.13-2.15). Stratified cumulative in-hospital mortality rates are given in the Supplement (Table 8).

**Table 1.**
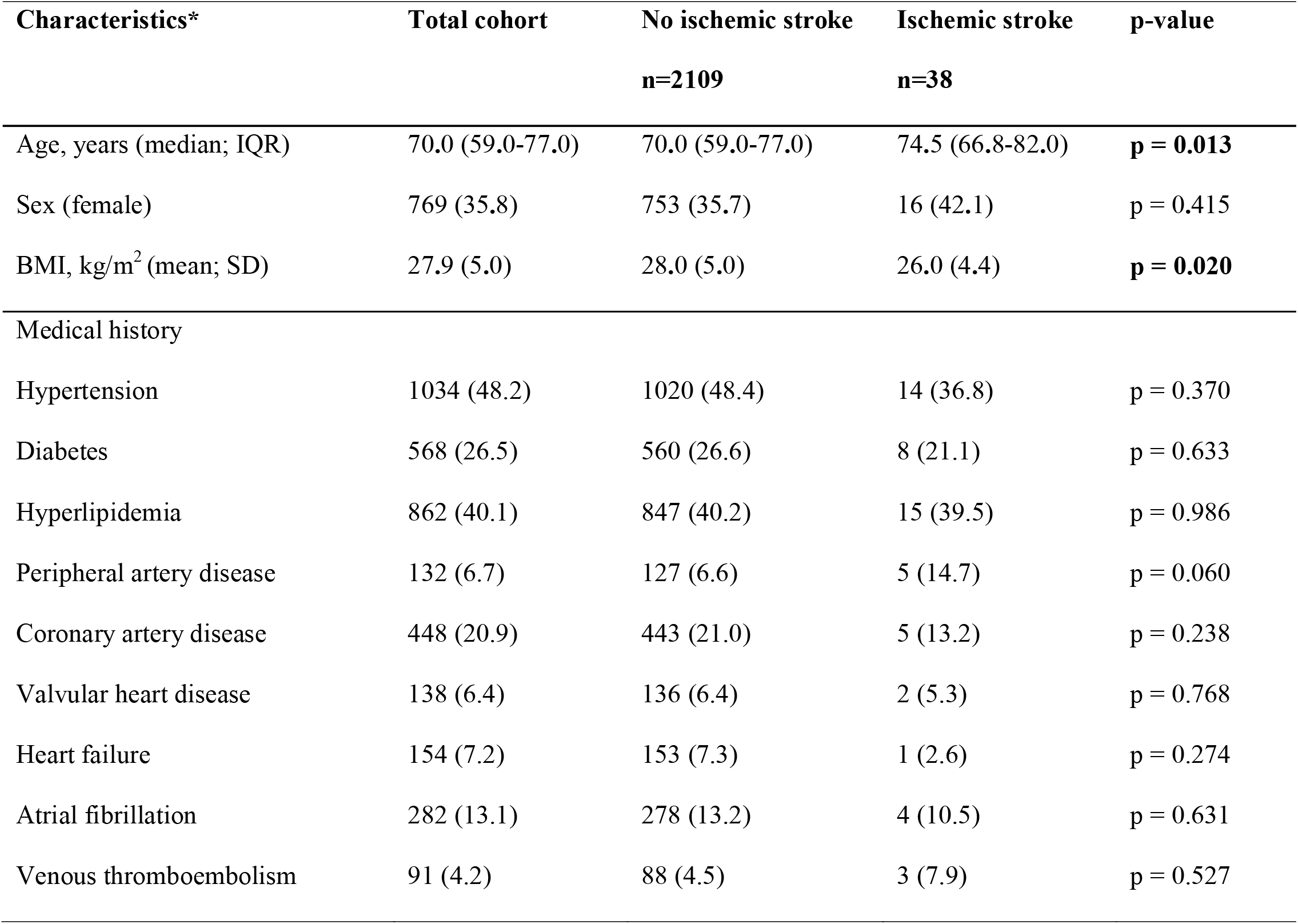
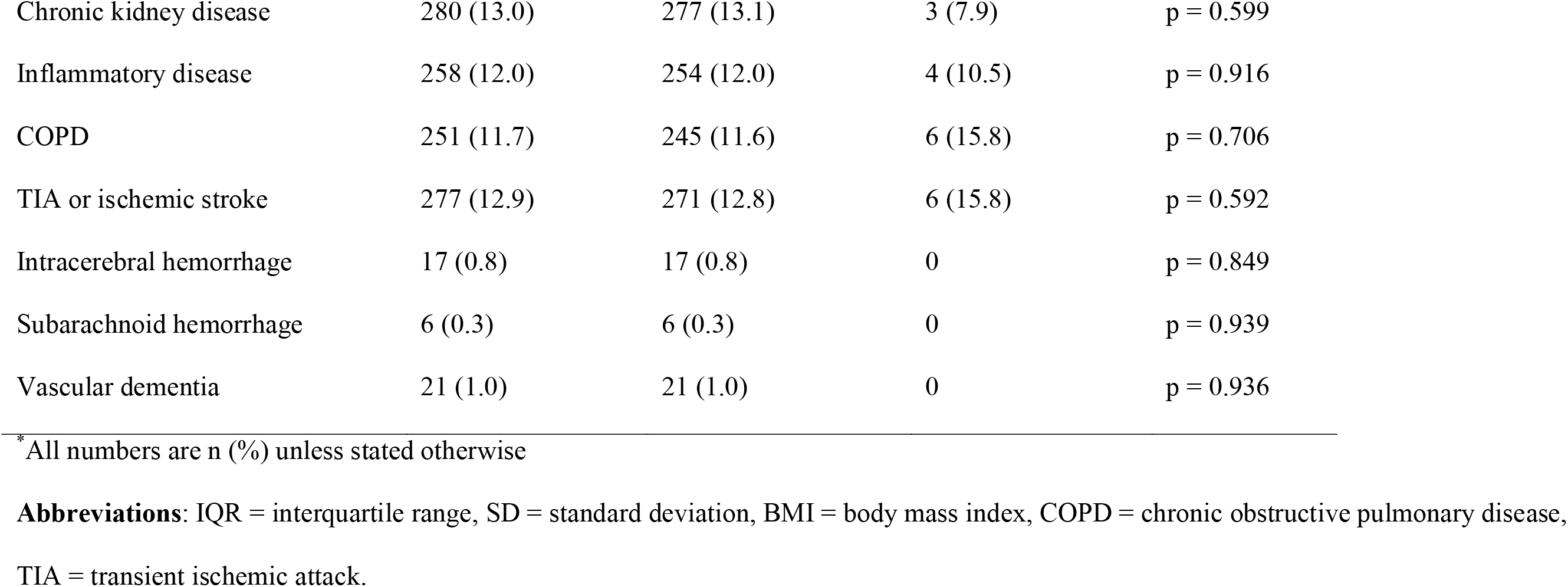
Baseline characteristics of hospitalized patients with COVID-19, stratified by diagnosis of acute ischemic stroke.

**Table 2.** Cumulative incidence of acute ischemic stroke in patients with and without treatment at an ICU, stratified by age and sex.

|  | **Total cohort** | **ICU treatment** | **General ward** |
| --- | --- | --- | --- |
|  | **(%; 95%CI)** | **(%; 95%CI)** | **(%; 95%CI)** |
| All ischemic stroke patients | 38/2147 (1.8; 1.3-2.4) | 16/586 (2.7; 1.7-4.4) | 22/1561 (1.4; 0.9-2.1) |
| Stratified by age |  |  |  |
| <50 years | 2/204 (1.0; 0.3-3.5) | 2/49 (4.1; 1.1-13.7) | 0/155 (0.0; 0-2.4) |
| 50-69 years | 11/816 (1.3; 0.8-3.5) | 9/318 (2.8; 1.5-5.3) | 2/498 (0.4; 0.1-1.5) |
| ≥70 years | 25/1127 (2.2; 1.5-3.3) | 5/219 (2.3; 1.0-5.2) | 20/908 (2.2; 1.4-3.4) |
| Stratified by sex |  |  |  |
| Female | 16/769 (2.1; 1.3-3.4) | 6/157 (3.8; 1.8-8.1) | 10/612 (1.6; 0.9-3.0) |
| Male | 22/1378 (1.6; 1.1-2.5) | 10/429 (2.3; 1.3-4.2) | 12/949 (1.3; 0.7-2.2) |

**Abbreviations:** CI = confidence interval.

**Table 3.**
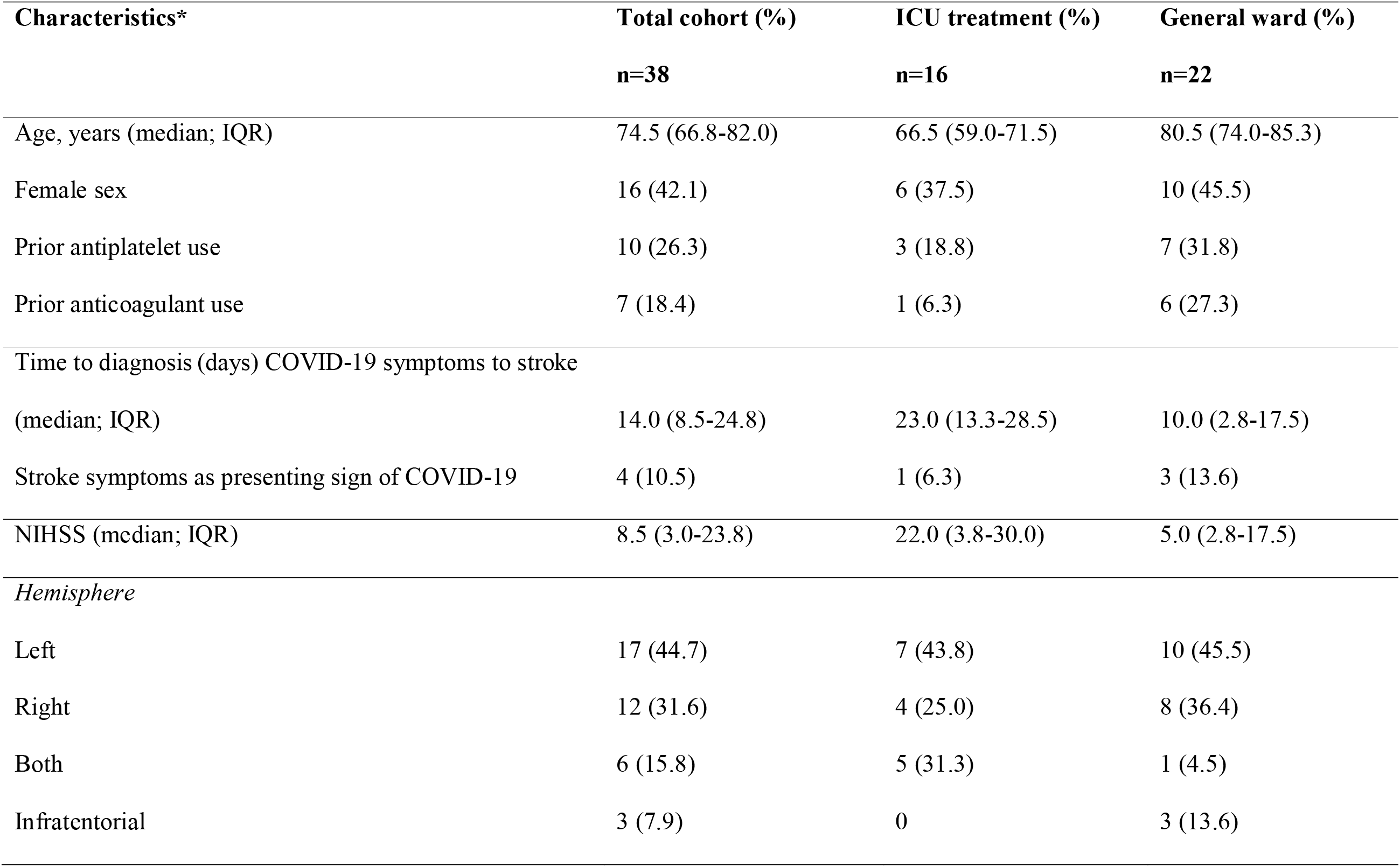
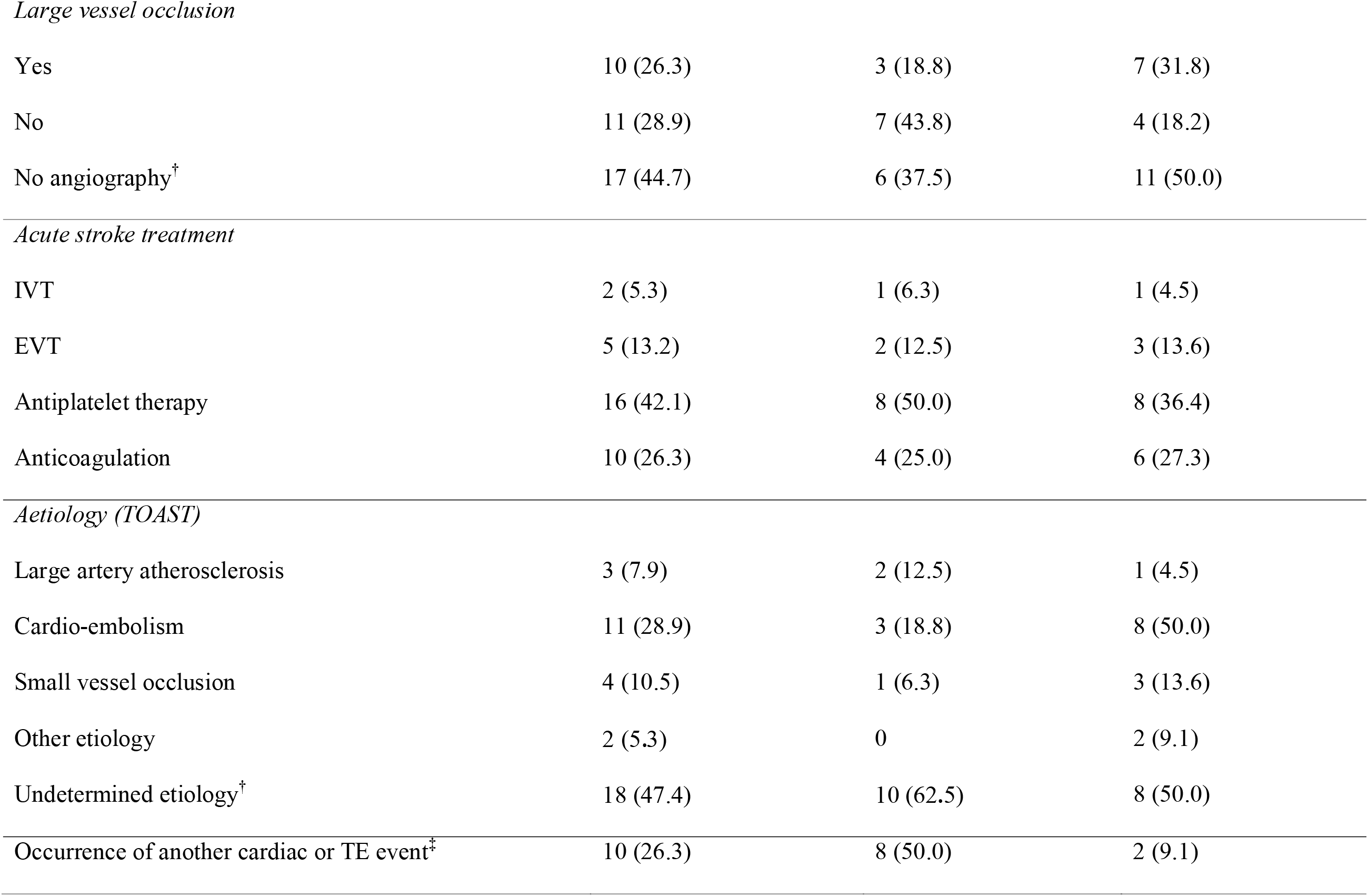
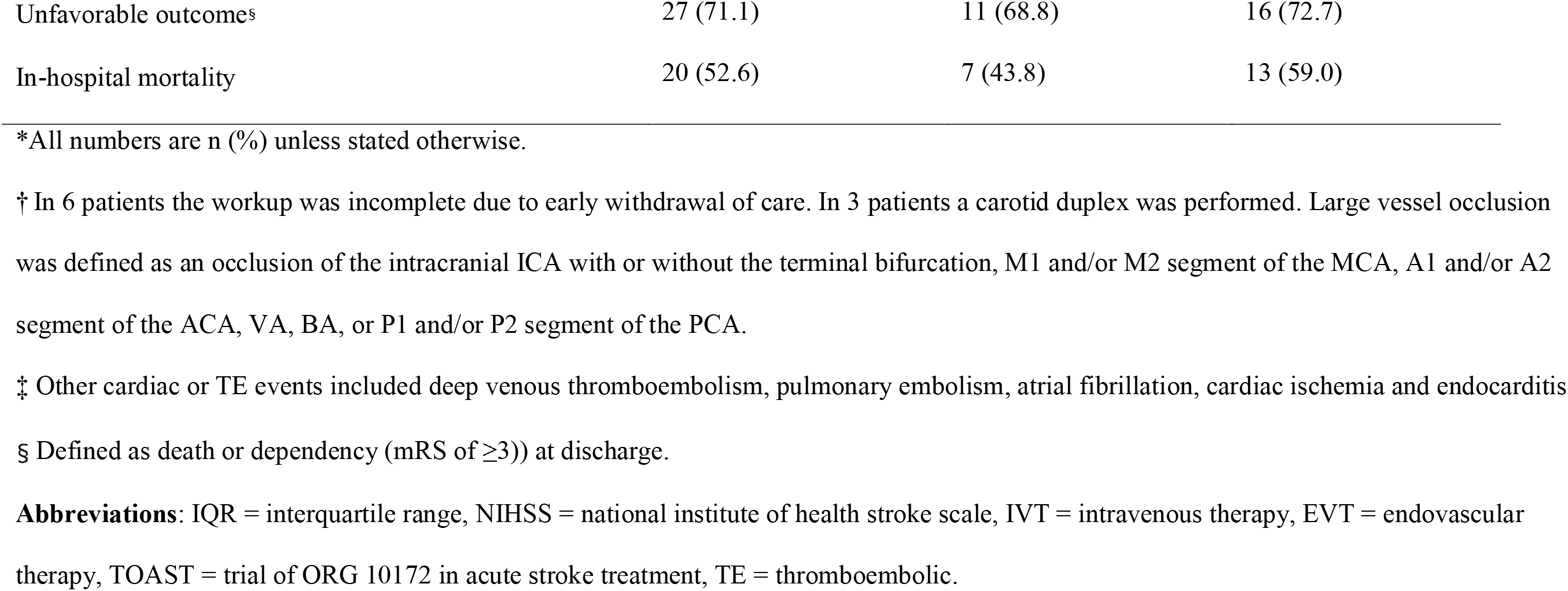
Characteristics of acute ischemic stroke in patients with COVID-19 treated at an ICU or on a general ward.

| **Characteristics\*** | **Total cohort (%)** | **ICU treatment (%)** | **General ward (%)** |
| --- | --- | --- | --- |
|  | **n=38** | **n=16** | **n=22** |
| Age, years (median; IQR) | 74.5 (66.8-82.0) | 66.5 (59.0-71.5) | 80.5 (74.0-85.3) |
| Female sex | 16 (42.1) | 6 (37.5) | 10 (45.5) |
| Prior antiplatelet use | 10 (26.3) | 3 (18.8) | 7 (31.8) |
| Prior anticoagulant use | 7 (18.4) | 1 (6.3) | 6 (27.3) |
| Time to diagnosis (days) COVID-19 symptoms to stroke |  |  |  |
| (median; IQR) | 14.0 (8.5-24.8) | 23.0 (13.3-28.5) | 10.0 (2.8-17.5) |
| Stroke symptoms as presenting sign of COVID-19 | 4 (10.5) | 1 (6.3) | 3 (13.6) |
| NIHSS (median; IQR) | 8.5 (3.0-23.8) | 22.0 (3.8-30.0) | 5.0 (2.8-17.5) |
| *Hemisphere* |  |  |  |
| Left | 17 (44.7) | 7 (43.8) | 10 (45.5) |
| Right | 12 (31.6) | 4 (25.0) | 8 (36.4) |
| Both | 6 (15.8) | 5 (31.3) | 1 (4.5) |
| Infratentorial | 3 (7.9) | 0 | 3 (13.6) |
| *Large vessel occlusion* |  |  |  |
| Yes | 10 (26.3) | 3 (18.8) | 7 (31.8) |
| No | 11 (28.9) | 7 (43.8) | 4 (18.2) |
| No angiography<sup>†</sup> | 17 (44.7) | 6 (37.5) | 11 (50.0) |
| *Acute stroke treatment* |  |  |  |
| IVT | 2 (5.3) | 1 (6.3) | 1 (4.5) |
| EVT | 5 (13.2) | 2 (12.5) | 3 (13.6) |
| Antiplatelet therapy | 16 (42.1) | 8 (50.0) | 8 (36.4) |
| Anticoagulation | 10 (26.3) | 4 (25.0) | 6 (27.3) |
| *Aetiology (TOAST)* |  |  |  |
| Large artery atherosclerosis | 3 (7.9) | 2 (12.5) | 1 (4.5) |
| Cardio-embolism | 11 (28.9) | 3 (18.8) | 8 (50.0) |
| Small vessel occlusion | 4 (10.5) | 1 (6.3) | 3 (13.6) |
| Other etiology | 2 (5.3) | 0 | 2 (9.1) |
| Undetermined etiology<sup>†</sup> | 18 (47.4) | 10 (62.5) | 8 (50.0) |
| Occurrence of another cardiac or TE event<sup>‡</sup> | 10 (26.3) | 8 (50.0) | 2 (9.1) |
| Unfavorable outcome<sup>§</sup> | 27 (71.1) | 11 (68.8) | 16 (72.7) |
| In-hospital mortality | 20 (52.6) | 7 (43.8) | 13 (59.0) |

\*All numbers are n (%) unless stated otherwise.
† In 6 patients the workup was incomplete due to early withdrawal of care. In 3 patients a carotid duplex was performed. Large vessel occlusion was defined as an occlusion of the intracranial ICA with or without the terminal bifurcation, M1 and/or M2 segment of the MCA, A1 and/or A2 segment of the ACA, VA, BA, or P1 and/or P2 segment of the PCA.
‡ Other cardiac or TE events included deep venous thromboembolism, pulmonary embolism, atrial fibrillation, cardiac ischemia and endocarditis
§ Defined as death or dependency (mRS of $\geq 3$ ) at discharge.
**Abbreviations:** IQR = interquartile range, NIHSS = national institute of health stroke scale, IVT = intravenous therapy, EVT = endovascular therapy, TOAST = trial of ORG 10172 in acute stroke treatment, TE = thromboembolic.

**Figure 1.**
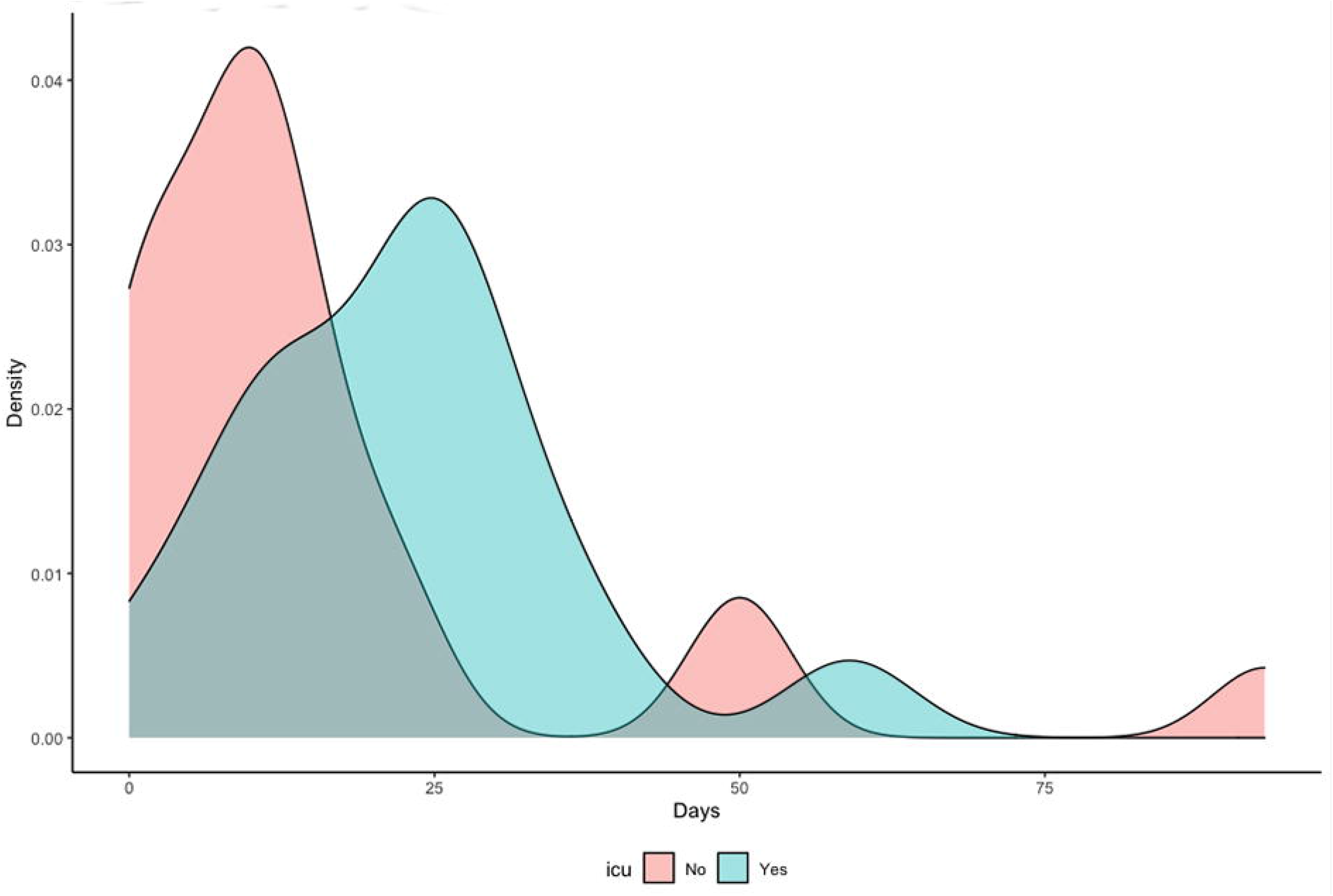
Median time between onset of COVID-19 symptoms and diagnosis of acute ischemic stroke in patients treated at an ICU or on a general ward.

**Figure 2.**
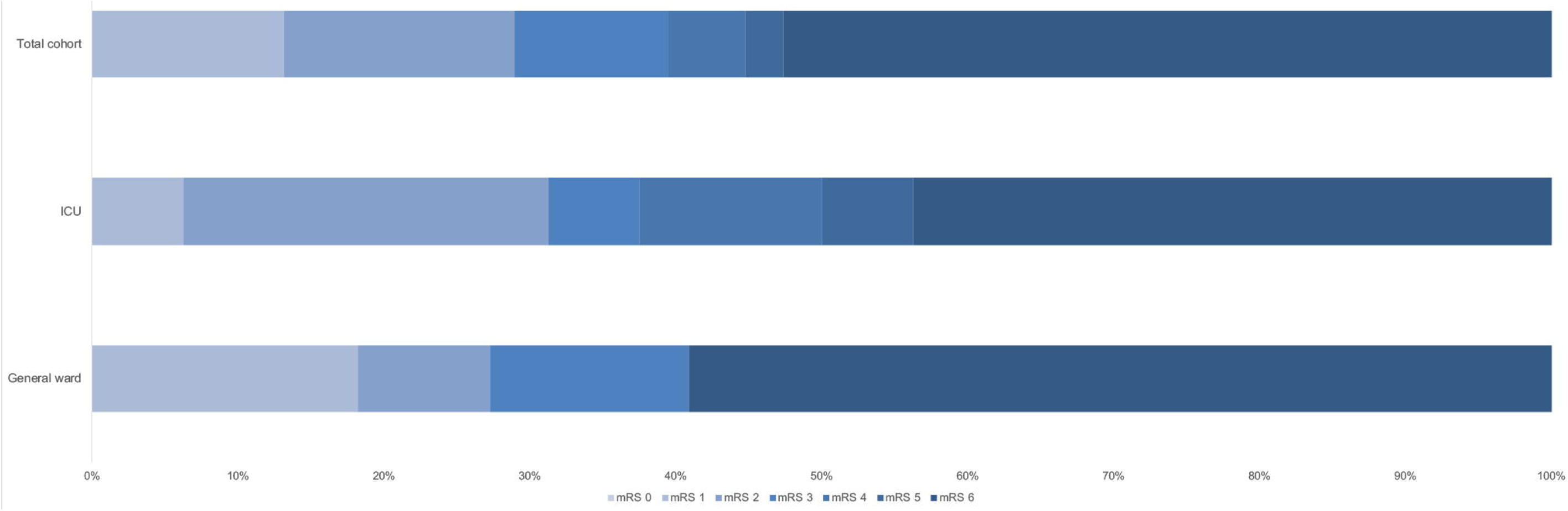
Outcome of acute ischemic stroke in patients with COVID-19 assessed with the modified Rankin Scale at discharge in patients with and without treatment at an ICU. **Abbreviations:** ICU = Intensive Care Unit, mRS = modified Rankin Scale.

## Discussion

In our multicenter cohort of hospitalized patients with COVID-19, the overall cumulative incidence of acute ischemic stroke was 1.8%, with a higher rate of ischemic stroke in patients who needed treatment at an ICU (2.7%). Patients with acute ischemic stroke were older, but did not have more cardiovascular risk factors. In addition, patients with ischemic stroke were twice as likely to have pulmonary embolism and were at higher risk of in-hospital mortality.

The overall incidence of ischemic stroke of 1.8% in hospitalized patients with COVID-19 is in line with previously published hospital-based COVID-19 cohorts, which reported cumulative incidences ranging between 1.0% and 2.4%.^3-5^ Lower stroke rates have been found in studies that reported on a combination of hospitalized and non-hospitalized patients.^6, 23^ Higher rates of up to 6.9% have been reported in populations consisting of ICU patients alone or in other selected patient populations.^5, 24, 25^ In addition to differences in population and hospital setting, the variation in reported incidence of ischemic stroke may also be explained by other factors. First, most studies were performed in Asia and North America, with only a few European cohorts.^3-5, 7^ Geographical variation may explain some of the heterogeneity, with a higher incidence reported in Asia than in North America and Europe.^3^ In addition, regional differences in COVID-19 surges may have resulted in a higher threshold for seeking medical attention in some pandemic areas, especially for patients with mild symptoms of stroke. Second, in most studies ischemic stroke was recorded as one of various cardiovascular events, with case-ascertainment often not performed by neurologists or stroke physicians.^6-8, 26^ This may have resulted in a systematic bias in the estimation of the cumulative stroke incidence among hospitalized patients with COVID-19.

In contrast to some of the previous cohorts,^24, 27^ our findings suggest that patients with COVID-19 who had acute ischemic stroke did not have more cardiovascular risk factors than patients who did not have a stroke. One explanation for this discrepancy may be that older patients with more vascular risk factors may not have been hospitalized or admitted to an ICU in the first place, because of treatment restrictions or patient preferences, which may have led to reduced survival rates in this group.^28^ In addition, the greater severity of COVID-19 illness among hospitalized patients, especially those treated at an ICU, as well the increased risk of medical complications during hospitalization, such as venous thromboembolism due to immobilisation, may, at least partially, have contributed to the stroke risk in hospitalized COVID-19 patients without vascular risk factors.^29^

To our knowledge, this is the first study to date to report on an association between pulmonary embolism and acute ischemic stroke in hospitalized patients with COVID-19 and to illustrate the time course of these cardiovascular complications. Although these findings need to be confirmed in additional studies, they may shed new light on the mechanisms by which COVID-19 is related to ischemic stroke, particularly in patients with no other vascular risk factors.

Acute respiratory infections in general can act as a trigger for the short-term risk of ischemic stroke and myocardial infarction and are associated with a high risk of cardiovascular-related death, especially in older patients and in those with cardiovascular comorbidity.^30, 31^ Two recent studies have compared the occurrence of acute ischemic stroke in hospitalized patients with COVID-19 versus those with influenza. One study found that patients with COVID-19 appeared to have an increased stroke risk (COVID-19: 1.6%; influenza: 0.2%), whereas the other study found the risk of ischemic stroke to be similar in patients with COVID-19 (1.2%) and influenza (1.2%).^9, 27^ In SARS-CoV (SARS) and Middle-East Respiratory Syndrome (MERS), the occurrence of acute ischemic stroke has only been reported sporadically.^32^ Pathophysiological mechanisms that could link COVID-19 to thromboembolic events include direct viral-induced endotheliitis, postinfectious immune-mediated responses, prothrombotic coagulopathy, and the occurrence of a hyperinflammatory state, with elevated D-dimer levels and antiphospholipid antibodies frequently found in patients with COVID-19 and thromboembolic complications.^2^

Several studies have found that patients with COVID-19 who had acute ischemic stroke were more likely to die.^3-5^ It remains unclear whether this association with an increased mortality is driven by disease severity and the prothrombotic state triggered by COVID-19. Alternatively, other confounding factors, such as impeded functional recovery due to fever and infection and withdrawal of care in patients with COVID-19 and ischemic stroke, may also play a role.^33, 34^

Our study has some limitations. First, different forms of bias should be considered in observational research. Hospitalized patients with COVID-19, and in particular those requiring treatment at an ICU, represent a selected group. Numerous factors may have influenced whether patients sought emergency care, were admitted to a hospital and received intensive treatment. Some patients with COVID-19 and ischemic stroke may have died before reaching the hospital, but alternatively milder affected patients or those with treatment restrictions may have stayed at home.^35^ This may have underestimated the overall rate of ischemic stroke in patients hospitalized with COVID-19. The high caseload of COVID-19 patients in some hospitals, in combination with contagion containment and sedation on an ICU, may have impeded imaging investigations to diagnose ischemic strokes, especially among moribund patients. This may have resulted in an overestimation of the percentage of strokes with undetermined etiology in our study and others. Among patients with pulmonary embolism and ischemic stroke, the diagnostic work-up to rule out a patent foramen ovale was often not performed. In contrast, the relatively large proportion of patients with a cardioembolic atiology may reflect the accessibility of telemetry and ECG. Second, we were unable to adjust for changes in admission and treatment strategies that occurred during the first wave of the pandemic. This may have affected the rate of ischemic stroke and in-hospital mortality and may hamper the generalizability of our results to later waves and other settings. Due to the novelty of this pandemic, comparisons with hospital populations from previous years and across different waves of COVID-19 should be interpreted with caution.^7, 36^ The main overall strength of the CAPACITY-COVID consortium is that it is a multinational and multi-disciplinary collaborative effort to systematically record all cardiovascular complications in patients with COVID-19 in a longitudinal fashion. By extending the consortium with STROCORONA, we were able not only to add cerebrovascular expertise and detailed acute ischemic stroke data, it also allowed us to link various cardiovascular complications in hospitalized patients with COVID-19.

## Conclusion

In conclusion, in our multicenter cohort the overall cumulative incidence of acute ischemic stroke in hospitalized patients with COVID-19 was approximately 2%, with a higher risk in patients treated at an ICU. Our finding that patients with COVID-19 and ischemic stroke were twice as likely to have pulmonary embolism than patients without stroke warrants further investigation, but may be relevant for our understanding of thrombotic mechanisms associated with COVID-19. Irrespective of potential pathophysiological relevance, our findings underscore the importance of appropriate antithrombotic strategies and increased awareness of stroke symptoms in hospitalized patients with COVID-19.

## Data Availability

The data used for this study, including de-identified individual participant data and a data dictionary defining each field or variable within the dataset, can be made available upon reasonable request to the data access committee of CAPACITY-COVID. These data will be made available following publication of this work. A data sharing agreement must be signed before any data are shared.

## Acknowledgements

We want to express our gratitude to all participating sites and researchers participating in the CAPACITY-COVID collaborative consortium. A list of all participating organizations is given in the Supplement (Table 9).

## Disclosures

Wouter M. Sluis is supported by the European Union’s Horizon, 2020 research and innovation programme (grant no. 634809).

Marijke Linschoten is supported by the Alexandre Suerman Stipend of the University Medical Center Utrecht.

J.P. Kerklaan participates in the PACIFIC-stroke trial.

Folkert W. Asselbergs is supported by University College London Hospitals National Institute for Health Research Biomedical Research and CardioVasculair Onderzoek Nederland 2015-12 eDETECT.

M.J.H Wermer is supported by a personal grant from the Dutch Heart Foundation (Dr. Dekker Grant 2016T086), a personal VIDI grant from ZonMw/NWO (91717337) and a grant for the CORONIS project from ZonMW and the Dutch Heart Foundation. She served as a consultant to Biogen without payment.

H.B. van der Worp served as a consultant to Bayer and LivaNova, with fees paid to his institution.

Annemijn M. Algra is supported by a personal grant from the Dutch Heart Foundation (Dr. Dekker Grant 2016T023).

## Abbreviations

COVID-19: Coronavirus disease 2019
ICU: Intensive Care Unit
PCR: Polymerase chain reaction
PE: Pulmonary embolism
AF: Atrial fibrillation
TIA: Transient ischemic attack
NIHSS: National Institutes of Health Stroke Scale
IVT: Intravenous thrombolysis
EVT: Endovascular therapy
TOAST: Trial of Org 10172 in Acute Stroke Treatment
mRS: Modified Rankin scale
IQR: Interquartile range
SD: Standard deviation
CI: Confidence interval
RR: Risk ratio

## Appendix

### The CAPACITY-COVID collaborative consortium

Aaf F.M. Kuijper MD PhD^1^, Clara E.E. van Ofwegen-Hanekamp MD PhD^2^, Richard C.J.M. Donders MD PhD^3^, D. Martijn O. Pruijsen MD PhD^3^, Rik S. Hermanides MD PhD^4^, Hortence, E. Haerkens-Arends MD^5^, Rutger L. Anthonio MD^6^, Mireille E. Emans MD PhD^7^, René A. Tio MD PhD^8^, Jur M. ten Berg MD PhD^9,10^, Björn E. Groenemeijer MD PhD^11^, Ron Pisters MD PhD^12^, P. Marc van der Zee MD PhD^13^, Hans-Marc J. Siebelink MD PhD^14^ Derk O. Verschure MD PhD^15^, Matthijs F.L. Meijs MD PhD^16^

1. Department of Cardiology, Spaarne Gasthuis, Haarlem, The Netherlands

2. Department of Cardiology, Diakonessenhuis Utrecht, Utrecht, The Netherlands

3. Department of Cardiology, Isala Hospital, Zwolle, The Netherlands

4 Department of Neurology, Diakonessenhuis Hospital, Utrecht, the Netherlands

5. Department of Cardiology, Jeroen Bosch Hospital, ‘s-Hertogenbosch, The Netherlands

6. Department of Cardiology, Treant Zorggroep, Emmen, The Netherlands

7. Department of Cardiology, Ikazia Hospital, Rotterdam, The Netherlands

8. Department of Cardiology, Catharina Hospital, Eindhoven, the Netherlands

9. Department of Educational Development and Research, Faculty of Health, Medicine and Life Sciences, Maastricht University, Maastricht, The Netherlands

10. Department of Cardiology, St. Antonius Hospital, Nieuwegein, the Netherlands

11. Department of Cardiology, Gelre Hospital Apeldoorn, Apeldoorn, The Netherlands

12. Department of Cardiology, Rijnstate Hospital, Arnhem, The Netherlands

13. Department of Cardiology, St. Jansdal Hospital, Harderwijk, the Netherlands

14. Department of Cardiology, HeartLungCenter, Leiden University Medical Center, Leiden, The Netherlands

15. Department of Cardiology, Zaans Medical Center, Zaandam, The Netherlands

16. Deparment of Cardiology, Medisch Spectrum Twente, Enschede, The Netherlands

