## Supplementary Table for "Risk, clinical course and outcome of ischemic stroke in patients hospitalized with COVID-19: a multicenter cohort study"


| | Title | Page |
|---|---|---|
| **Figure 1** | Flowchart of patient inclusion | 2 |
| **Table 1** | Missing data | 3 |
| **Table 2** | Baseline characteristics stratified according to the need of ICU treatment | 6 |
| **Table 3** | Baseline medication in patients with and without acute ischemic stroke | 8 |
| **Table 4** | Baseline characteristics stratified according to age groups | 10 |
| **Table 5** | Sensitivity analysis excluding centers that included patients with cardiovascular risk factors only | 12 |
| **Table 6** | Occurrence of other thromboembolic complications in patients with and without acute ischemic stroke stratified by etiology of stroke | 14 |
| **Table 7** | Occurrence of other thromboembolic complications in patients with and without acute ischemic stroke stratified by age and sex | 16 |
| **Table 8** | In-hospital mortality in patients with COVID-19 with an ischemic stroke according to the need of ICU treatment and stratified by age and sex | 17 |
| **Table 9** | List of participating organizations in CAPACITY COVID | 18 |

**Figure 1.** Flowchart of patient inclusion

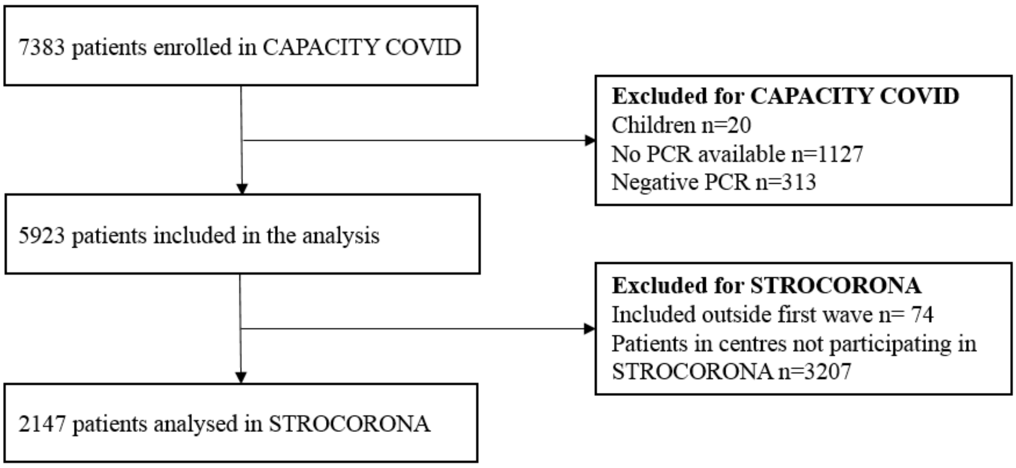

**Table 1.** Missing data

| Characteristics | Missing values (%) |
|---|---|
| Age, years (mean; SD) | 0 |
| Sex (female) | 0 |
| BMI, kg/m$^2$ (mean; SD) | 359 (16.7) |
| Ethnicity | 358 (16.7) |
| **Medical history** | |
| Hypertension | 48 (2.2) |
| Diabetes | 16 (0.7) |
| Hyperlipidemia | 181 (8.4) |
| Peripheral artery disease | 179 (8.3) |
| Coronary artery disease | 184 (8.6) |
| Valvular heart disease | 0 |
| Heart failure | 0 |
| Atrial fibrillation | 0 |
| Venous thromboembolism | 184 (8.6) |
| Chronic kidney disease | 6 (0.3) |
| Inflammatory disease | 5 (0.2) |
| COPD | 4 (0.2) |
| TIA and/or Ischemic stroke | 0 |
| Intracerebral hemorrhage | 1 (<0.1) |
| Subarachnoid hemorrhage | 1 (<0.1) |

| | |
|---|---|
| **Baseline medication** | |
| Betablockers | 3 (0.1) |
| Anti-arrhythmic drugs | 3 (0.1) |
| Digoxin | 3 (0.1) |
| Diuretics | 3 (0.1) |
| Calciumchannel blockers | 3 (0.1) |
| ACE inhibitors | 3 (0.1) |
| Angiotensin receptor blocker | 3 (0.1) |
| Aldosteron antagonists | 3 (0.1) |
| Coumarins | 3 (0.1) |
| DOAC | 3 (0.1) |
| Lipid lowering medication | 3 (0.1) |
| Insulin | 3 (0.1) |
| Oral anti diabetics | 3 (0.1) |
| Anti-platelet therapy* | 5 (0.2) |
| Dual platelet therapy† | 2 (0.3) |
| **In-hospital complications** | |
| Deep venous thromboembolism | 0 |
| Pulmonary embolism | 0 |
| Atrial fibrillation | 0 |
| Cardiac ischaemia | 0 |
| Endocarditis | 0 |
| Acute ischemic stroke | 0 |
| In-hospital mortality | 0 |

* usage of anti-platelet therapy in general. † usage of more than one anti-platelet drug.

**Abbreviations**: SD = standard deviation, BMI = body mass index, COPD = chronic obstructive pulmonary disease, TIA = transient ischemic attack, ACE = angiotensin converting enzyme, DOAC = direct oral anticoagulant, AIS = acute ischemic stroke.

**Table 2.** Baseline characteristics stratified according to the need of ICU treatment

| **Characteristics*** | **ICU treatment**<br>**n=586** | **No ICU treatment**<br>**n=1561** | **p-value** |
|---|---|---|---|
| Age, years (median; IQR) | 66.0 (58.0-72.3) | 73.0 (60.0-80.0) | **p = <0.001** |
| Sex (female) | 157 (26.8) | 612 (39.2) | **p = <0.001** |
| BMI, kg/m$^2$ (mean; SD) | 28.5 (4.8) | 27.7 (5.1) | **p = 0.002** |
| **Medical history** | | | |
| Hypertension | 240 (41.0) | 794 (50.9) | **p = <0.001** |
| Diabetes | 141 (24.1) | 427 (27.4) | p = 0.293 |
| Hyperlipidemia | 200 (34.1) | 662 (42.4) | **p = <0.001** |
| Peripheral artery disease | 32 (5.8) | 100 (7.1) | p = 0.301 |
| Coronary artery disease | 95 (16.2) | 353 (22.6) | **p = 0.001** |
| Valvular heart disease | 18 (3.1) | 120 (7.7) | **p = <0.001** |
| Heart failure | 17 (2.9) | 137 (8.8) | **p = <0.001** |
| Atrial fibrillation | 52 (8.9) | 230 (14.7) | **p = <0.001** |
| Venous thromboembolism | 20 (3.4) | 71 (4.5) | **p = <0.001** |
| Chronic kidney disease | 44 (7.5) | 236 (15.1) | **p = <0.001** |
| Inflammatory disease | 48 (8.2) | 210 (13.5) | **p = 0.003** |
| COPD | 47 (8.0) | 204 (13.1) | **p = 0.005** |
| TIA and/or Ischemic stroke | 52 (8.9) | 225 (14.4) | **p = <0.001** |
| Intracerebral hemorrhage | 4 (0.7) | 13 (0.8) | p = 0.248 |
| Subarachnoid hemorrhage | 2 (0.3) | 4 (0.3) | p = 0.249 |

*Numbers are n (%) unless otherwise stated.

**Abbreviations:** ICU = critical or intensive care unit, IQR = interquartile range, SD = standard deviation, BMI = body mass index, COPD = chronic obstructive pulmonary disease, TIA = transient ischemic attack.

**Table 3.** Baseline medication in patients with and without acute ischemic stroke

| Baseline medication* | Total cohort<br>n=2147 | No ischemic stroke<br>n=2109 | Ischemic stroke<br>n=38 | p-value |
|---|---|---|---|---|
| Betablockers | 653 (30.4) | 642 (30.4) | 11 (28.9) | p = 0.953 |
| Anti-arrhythmic drugs | 91 (4.2) | 90 (4.3) | 1 (2.6) | p = 0.860 |
| Digoxin | 47 (2.2) | 47 (2.2) | 0 | p = 0.631 |
| Diuretics | 523 (24.4) | 515 (24.4) | 8 (21.1) | p = 0.866 |
| Calciumchannel blockers | 348 (16.2) | 345 (16.4) | 3 (7.9) | p = 0.362 |
| ACE inhibitors | 422 (19.7) | 416 (19.7) | 6 (15.8) | p = 0.808 |
| Angiotensin receptor blocker | 312 (14.5) | 306 (14.5) | 6 (15.8) | p = 0.950 |
| Aldosteron antagonists | 71 (3.3) | 71 (3.4) | 0 | p = 0.501 |
| Coumarins | 163 (7.6) | 160 (7.6) | 3 (7.9) | p = 0.971 |
| DOAC | 201 (9.4) | 197 (9.3) | 4 (10.5) | p = 0.944 |
| Lipid lowering medication | 839 (39.1) | 823 (39.0) | 16 (42.1) | p = 0.906 |
| Insulin | 183 (8.5) | 181 (8.6) | 2 (5.3) | p = 0.746 |
| Oral anti diabetics | 377 (17.6) | 371 (17.6) | 6 (15.8) | p = 0.932 |

| | | | | |
|---|---|---|---|---|
| Anti-platelet therapy | 639 (29.8) | 629 (29.8) | 10 (26.3) | p = 0.852 |
| Dual platelet therapy | 57 (8.9) | 55 (8.8) | 2 (20.0) | p = 0.217 |

*Numbers are n (%).

**Abbreviations:** ACE = angiotensin converting enzyme, DOAC = direct oral anticoagulant**.**

**Table 4.** Baseline characteristics stratified according to age groups

| Characteristics* | Age <50 years | | Age 50-69 years | | Age ≥70 years | |
|---|---|---|---|---|---|---|
| | No ischemic stroke<br>n= 202 | Ischemic stroke<br>n= 2 | No ischemic stroke<br>n= 805 | Ischemic stroke<br>n= 11 | No ischemic stroke<br>n= 1102 | Ischemic stroke<br>n= 25 |
| **Medical history** | | | | | | |
| Hypertension | 35 (17.3) | 0 | 312 (38.8) | 3 (27.3) | 673 (61.1) | 11 (44.0) |
| Diabetes | 33 (16.3) | 1 (50.0) | 179 (22.2) | 0 | 348 (31.6) | 7 (28.0) |
| Hyperlipidemia | 22 (10.9) | 1 (50.0) | 267 (33.2) | 5 (45.5) | 558 (50.6) | 9 (36.0) |
| Peripheral artery disease | 6 (3.1) | 0 | 30 (4.0) | 1 (12.5) | 91 (9.3) | 4 (16.7) |
| Coronary artery disease | 4 (2.0) | 0 | 117 (14.5) | 1 (9.1) | 322 (29.2) | 4 (16.0) |
| Valvular heart disease | 0 | 0 | 18 (2.2) | 0 | 118 (10.7) | 2 (8.0) |
| Heart failure | 1 (0.5) | 0 | 22 (2.7) | 0 | 130 (11.8) | 1 (4.0) |
| Atrial fibrillation | 1 (0.5) | 0 | 38 (4.7) | 0 | 239 (21.7) | 4 (16.0) |
| Venous embolism | 2 (1.0) | 0 | 29 (3.6) | 0 | 57 (5.2) | 3 (12.0) |
| Chronic kidney disease | 12 (5.9) | 0 | 52 (6.5) | 1 (9.1) | 213 (19.3) | 2 (8.0) |

| | | | | | | |
|---|---|---|---|---|---|---|
| Inflammatory disease | 23 (11.4) | 0 | 91 (11.3) | 1 (9.1) | 140 (12.7) | 3 (12.0) |
| COPD | 3 (1.5) | 0 | 71 (8.8) | 2 (18.2) | 171 (15.5) | 4 (16.0) |
| TIA / Ischemic stroke | 4 (2.0) | 0 | 59 (7.3) | 2 (18.2) | 208 (18.9) | 4 (16.0) |
| Intracerebral hemorrhage | 0 | 0 | 6 (0.7) | 0 | 11 (1.0) | 0 |
| Subarachnoid hemorrhage | 0 | 0 | 3 (0.4) | 0 | 3 (0.3) | 0 |

*Numbers are n (%) unless otherwise stated.

**Abbreviations:** COPD = chronic obstructive pulmonary disease, TIA = transient ischemic attack.

**Table 5.** Sensitivity analysis excluding centers that included patients with cardiovascular risk factors only

| **Characteristics*** | **Total cohort** | **No ischemic stroke** | **Ischemic stroke** | **p-value** |
|---|---|---|---|---|
| n | 1846 | 1810 (98.0) | 36 (2.0) | |
| Age, years (median; IQR) | 70.0 (59.0-77.0) | 70.0 (59.0-77.0) | 74.0 (66.3-82.0) | **p = <0.001** |
| Sex (female) | 667 (36.1) | 653 (36.1) | 14 (38.9) | p = 0.728 |
| BMI, kg/m$^2$ (mean; SD) | 27.9 (5.1) | 27.9 (5.1) | 25.8 (4.4) | **p = 0.025** |
| **Medical history** | | | | |
| Hypertension | 852 (47.3) | 840 (47.6) | 12 (34.3) | p = 0.119 |
| Diabetes | 480 (26.2) | 473 (26.3) | 7 (19.4) | p = 0.354 |
| Hyperlipidemia | 721 (39.1) | 706 (39.0) | 15 (41.7) | p = 0.947 |
| Peripheral artery disease | 116 (6.5) | 111 (6.4) | 5 (15.2) | **p = 0.043** |
| Coronary artery disease | 352 (19.1) | 347 (19.2) | 5 (13.9) | p = 0.424 |
| Valvular heart disease | 112 (6.1) | 110 (6.1) | 2 (5.6) | p = 0.897 |
| Heart failure | 128 (6.9) | 128 (7.1) | 0 (0.0) | p = 0.098 |
| Atrial fibrillation | 242 (13.1) | 239 (13.2) | 3 (8.3) | p = 0.391 |
| Venous thromboembolism | 82 (4.4) | 79 (4.4) | 3 (8.3) | p = 0.456 |

| | | | | |
|---|---|---|---|---|
| Chronic kidney disease | 224 (12.1) | 222 (12.3) | 2 (5.6) | p = 0.469 |
| Inflammatory disease | 223 (12.1) | 220 (12.2) | 3 (8.3) | p = 0.776 |
| COPD | 212 (11.5) | 207 (11.4) | 5 (13.9) | p = 0.648 |
| TIA and/or Ischemic stroke | 239 (12.9) | 233 (12.9) | 6 (16.7) | p = 0.502 |
| Intracerebral hemorrhage | 15 (0.8) | 15 (0.8) | 0 (0.0) | p = 0.583 |
| Subarachnoid hemorrhage | 4 (0.2) | 4 (0.2) | 0 (0.0) | p = 0.778 |

*Numbers are n (%) unless otherwise stated.

**Abbreviations**: ICU = critical or intensive care unit, SD = standard deviation, BMI = body mass index, COPD = chronic obstructive pulmonary disease, TIA = transient ischemic attack.

**Table 6.** Occurrence of other thromboembolic complications in patients with and without acute ischemic stroke stratified by etiology

| **Characteristics*** | **Total cohort (%)**<br>**n=38** | **Cryptogenic (%)**<br>**n=18** | **Known etiology (%)**<br>**n=20** |
|---|---|---|---|
| Age, years (median; IQR) | 74.5 (66.8-82.0) | 71.5 (64.3-81.0) | 80.0 (67.0-82.0) |
| Female sex | 16 (42.1) | 8 (44.4) | 8 (40.0) |
| Prior antiplatelet use | 10 (26.3) | 6 (33.3) | 4 (20.0) |
| Prior anticoagulant use | 7 (18.4) | 0 (0.0) | 7 (35.0) |
| Time to diagnosis (median; IQR) | | | |
| Symptom to stroke[†] | 14.0 (7.5-26.3) | 23.0 (8.5-30.0) | 11.5 (7.5-15.8) |
| Stroke symptoms as presenting complaint | 4 (10.5) | 1 (5.5) | 3 (15.0) |
| Treatment at ICU | 16 (42.1) | 10 (55.6) | 6 (30.0) |
| NIHSS (median; IQR) | 8.5 (3.0-23.8) | 16.5 (4.5-30.0) | 5.0 (2.3-20.0) |
| *Hemisphere* | | | |
| Left | 17 (44.7) | 6 (33.3) | 11 (55.0) |
| Right | 12 (31.6) | 7 (38.9) | 5 (25.0) |
| Both | 6 (15.8) | 4 (22.2) | 2 (10.0) |

| | | | |
|---|---|---|---|
| Infratentorial | 3 (7.9) | 1 (5.6) | 2 (10.0) |
| *Large vessel occlusion* | | | |
| Yes | 10 (26.3) | 4 (22.2) | 6 (30.0) |
| No | 11 (28.9) | 5 (27.8) | 6 (30.0) |
| No angiography | 17 (44.7) | 9 (50.0) | 8 (40.0) |
| *Reperfusion therapy* | | | |
| IVT | 2 (5.3) | 1 (5.6) | 1 (5.0) |
| EVT | 5 (13.2) | 3 (16.7) | 2 (10.0) |
| Occurrence of another TE event | 10 (26.3) | 6 (33.3) | 4 (20.0) |
| Unfavorable outcome‡ | 27 (71.1) | 14 (77.8) | 13 (65.0) |
| In-hospital mortality | 20 (52.6) | 10 (55.6) | 10 (50.0) |

* All numbers are N (%) unless otherwise stated.

† Days from first COVID symptoms to diagnosis of ischemic stroke;

‡ Defined as death or dependency, modified Rankin scale 3 or higher

**Abbreviations**: IQR = interquartile range, NIHSS = national institute of health stroke scale, IVT = intravenous therapy, EVT = endovascular therapy, TOAST = trial of ORG 10172 in acute stroke treatment. TE=thromboembolic.

**Table 7.** Occurrence of other thromboembolic complications in patients with and without ischemic stroke stratified by age and sex

| | Total (n) | DVT (%) | | PE (%) | | AF (%) | | Cardiac ischemia (%) | | Endocarditis (%) | |
|---|---|---|---|---|---|---|---|---|---|---|---|
| | | No AIS | AIS | No AIS | AIS | No AIS | AIS | No AIS | AIS | No AIS | AIS |
| Overall | 2147 | 11 (0.5) | 0 | 160 (7.6) | 8 (21.1) | 145 (6.9) | 4 (10.5) | 36 (1.7) | 2 (5.3) | 2 (0.1) | 0 |
| Stratified by sex | | | | | | | | | | | |
| Females | 769 | 2 (0.3) | 0 | 35 (4.6) | 4 (25.0) | 44 (5.8) | 2 (12.5) | 7 (0.9) | 29 (2.1) | 1 (0.1) | 0 |
| Males | 1378 | 9 (0.7) | 0 | 125 (9.2) | 4 (18.2) | 101 (7.4) | 2 (9.1) | 29 (2.1) | 2 (9.1) | 1 (0.1) | 0 |
| Stratified by age | | | | | | | | | | | |
| <50 years | 204 | 1 (0.5) | 0 | 10 (5.0) | 1 (50.0) | 2 (1.0) | 0 | 0 | 0 | 0 | 0 |
| 50-69 years | 816 | 8 (1.0) | 0 | 93 (11.6) | 5 (45.5) | 53 (6.6) | 1 (9.1) | 18 (2.2) | 1 (9.1) | 1 (0.1) | 0 |
| ≥70 years | 1127 | 2 (0.2) | 0 | 57 (5.2) | 2 (8.0) | 90 (8.2) | 3 (12.0) | 18 (1.6) | 1 (4.0) | 1 (0.1) | 0 |

*Numbers are n (%).

**Abbreviations:** AIS = acute ischemic stroke, DVT = deep vein thrombosis, PE = pulmonary embolism, AF = atrial fibrillation.

**Table 8.** In-hospital mortality in patients with COVID-19 with an ischemic stroke according to the need of ICU treatment and stratified by age and sex

| | **Total cohort (%; 95%CI)** | **ICU treatment (%; 95%CI)** | **General ward (%; 95%CI)** |
|---|---|---|---|
| Total | 20/38 (52.6; 37.3-67.5) | 7/16 (43.8; 0.23-0.67) | 13/22 (59.1; 0.39-0.77) |
| Stratified by sex | | | |
| Female | 7/16 (43.8; 0.23-0.67) | 1/6 (16.7; 3.0-56.4) | 6/10 (60.0; 31.1-83.2) |
| Male | 13/22 (59.1; 0.39-0.77) | 6/10 (60.0; 31.1-83.2) | 7/12 (58.3; 32.0-80.7) |
| Stratified by age | | | |
| <50 years | 1/2 (50.0; 9.5-90.6) | 1/2 (50.0; 9.5-90.6) | 0 |
| 50-69 years | 4/11 (36.4; 15.2-64.6) | 3/9 (33.3; 12.1-64.6) | 1/2 (50.0; 9.5-90.6) |
| ≥70 years | 15/25 (60.0; 40.7-76.6) | 3/5 (60.0; 23.1-88.2) | 12/20 (60.0; 38.7-78.1) |

Numbers are n (%; 95%CI).

**Abbreviations:** CI = confidence interval

**Table 9.** List of participating and endorsing organizations in CAPACITY COVID

Dutch CardioVascular Alliance (DCVA)

The Dutch Association of Medical Specialists (FMS)

The British Heart Foundation Centres of Research Excellence.

The European Society of Cardiology (ESC)

The European Heart Network (EHN)

The Society for Cardiovascular Magnetic Resonance (SCMR)

The Dutch Society of Neurology

Kennisnetwerk CVA NL
